# Forgoing healthcare during the COVID-19 pandemic in Geneva, Switzerland – a cross-sectional population-based study

**DOI:** 10.1101/2021.10.01.21264411

**Authors:** Lakshmi Krishna Menon, Viviane Richard, Carlos de Mestral, Helene Baysson, Ania Wisniak, Idris Guessous, Silvia Stringhini, for the Specchio-COVID19 Study Group

## Abstract

**Background:** Health systems around the world continue to navigate through operational challenges surfaced by the COVID-19 pandemic; these have implications for access to healthcare. In this study, we estimate the prevalence and reasons for forgoing healthcare during the pandemic in Geneva, Switzerland; a country with a universal and mandatory private health insurance coverage.

**Methods:** Participants from a randomly selected population-based sample of the adult population living in the Canton of Geneva completed an online socio-demographic and lifestyle questionnaire between November 2020 and January 2021. The prevalence and reasons for forgoing healthcare since the beginning of the COVID-19 pandemic were examined descriptively, and logistic regression models were used to assess determinants for forgoing healthcare.

**Results:** The study included 5,397 participants, among which 8.0% reported having forgone healthcare since the beginning of the COVID-19 pandemic; participants with a disadvantaged financial situation (OR=2.04; 95% CI: 1.56-2.65), and those reporting an average (OR=2.55; 95% CI: 1.94-3.32) or poor health (OR=4.40; 95% CI: 2.40-7.68) were more likely to forgo healthcare. The most common reasons to forgo healthcare were appointment cancellations by healthcare providers (53.9%), fear of infection (35.3%), and personal organizational issues (11.1%).

**Conclusion:** Our paper highlights the effects of the COVID-19 pandemic on access to healthcare and identifies population sub-groups at-risk for forgoing healthcare. These results necessitate public health efforts to ensure equitable and accessible healthcare as the COVID-19 pandemic continues.

**Highlights:**

- 8% of the Geneva, Switzerland, adults renounced healthcare in the COVID-19 pandemic
- Main forgoing healthcare reasons are appointment cancellation and fear of infection
- Underprivileged participants with poor health are more likely to forgo healthcare

## Introduction

Owing to the rapid transmission of the SARS-CoV-2, responsible of the coronavirus disease 2019 (COVID-19), healthcare systems experienced major disruptions worldwide, including the temporary closure of medical practices, and cancellation or postponement of non-emergency and elective procedures [1,2]. These have induced individuals to forgo healthcare in preventive medicine, chronic disease diagnosis and management, and emergency medicine, as reported in several countries [1,3,4]. In Switzerland, a study conducted among vulnerable outpatients in the Geneva University Hospitals in June 2020 indicated that 38.5% had renounced healthcare since the start of the pandemic. [5]. However, this study relied on patient populations, whereas data from the general population remains limited. One such study in the United States showed that 41% of adults forwent healthcare between March and mid-June 2020 [6]. To date, no study has reported on the prevalence of forgoing healthcare during the COVID-19 pandemic in the general population of countries with universal healthcare coverage. This information is important to understand the extent to which the pandemic-related disruptions in the healthcare system may have impacted access to healthcare, particularly among vulnerable and socioeconomically underprivileged population groups. Indeed, forgoing or delaying healthcare may have health consequences [7]. For instance, forgoing healthcare for chronic and emergency conditions can lead to increased complications and costs [8,9]; while missing preventive care appointments, such as cancer screenings, can result in a delayed diagnosis and poorer prognosis [1]. In this paper, we used a population-based sample of the adult population living in Geneva, Switzerland to estimate the prevalence of forgoing healthcare during the COVID-19 pandemic and to assess the effect of anti-SARS-CoV-2 serological status, general health, and sociodemographic factors on the likelihood of forgoing healthcare.

## Methods

### Study Design

Our study population is part of SARS-CoV-2 serosurvey studies conducted in Geneva, Switzerland. Participants were randomly selected from this population at two periods. First, between April and June 2020, Geneva residents who were former participants of a yearly health survey representative of the general population, were invited for a free anti-SARS-CoV-2 serology, along with their family members [10]. Second, between November and December 2020, Geneva residents were randomly selected from a sex- and age-stratified cantonal register and invited to participate to a second serosurvey [11]. Adult participants from both studies were subsequently invited to participate in a longitudinal follow-up; the Specchio-COVID19 study [12]. The Specchio-COVID19 study was approved by the Cantonal Research Ethics Commission of Geneva, Switzerland (CCER Project ID 2020-00881) and informed consent was obtained from all participants. Participants completed a baseline questionnaire between November 2020 and January 2021 that collected demographic, socioeconomic, health and lifestyle information, including forgoing healthcare.

### Study Variables

All variables, apart from the SARS-CoV-2 serological status, were extracted from the Specchio-COVID19 baseline questionnaire. We defined forgoing healthcare as having answered “yes” to the question, “Since the start of the COVID-19 pandemic, have you renounced certain types of healthcare or treatment (by choice or constraint)?”. If the answer was “yes”, a following multiple-choice question collected information about the type of forgone healthcare. Options included: surgery; general practitioner; specialist; medication; dental; screening; inpatient rehabilitation; outpatient rehabilitation; medical devices; health center care; home nursing care; and other, from which we created three extra categories: “physiotherapy”, “mental care”, and “alternative care”, such as acupuncture or therapeutic massage. A third question collected information about the reason for forgoing healthcare: cancelled or postponed appointment by the healthcare provider due to the COVID-19 pandemic; fear of being infected when seeking healthcare; financial reasons; inability to go to the appointment because of quarantine, isolation, family or professional reasons; participant self-isolating due to possible SARS-CoV-2 exposure; and other, from which we created two more categories: “conscious”, for participants postponing healthcare to avoid overloading healthcare facilities, and “lethargy” for participants delaying healthcare that is unwanted. It was assumed that participants who did not report forgoing healthcare in the first place would not have selected any of these choice if they were asked to.

As potential risk factors, we included: country of birth, education (divided into *lower*, i.e. compulsory or no formal education; *medium* i.e. secondary education; and *higher* i.e. tertiary education), general health (derived from the question “In general, outside the pandemic context, how would you evaluate your health?”; answers were coded as *good* if “very good” or “good”, *medium* if “medium” and *poor* if “poor or very poor”), and financial situation (this was defined as *very good* if participants answered they could save money to the question “Currently, how would you assess your financial situation?”, *good* if they could afford minor unexpected expenses, and *average to poor* if they chose one of following statements; “I have to be careful with my expenses and an unexpected event could put me in financial difficulty” or “I cannot cover my needs with my income and I need external support”). We also included anti-SARS-CoV-2 serological status as covariate, measured as following. In the first serosurvey, seropositivity was detected with an enzyme-linked immunosorbent assay (ELISA) (Euroimmun, Lübeck, Germany #EI 2606– 9601 G). Positive and indeterminate (IgG ratio for detection ≥0.5) results were ascertained with a recombinant immunofluorescence assay (rIFA), as previously detailed [10]. For the second serosurvey, values ≥0·8 U/mL of the Elecsys anti-SARS-CoV-2 S (Roche Diagnostics, Rotkreuz, Switzerland) [13] were considered positive.

### Statistical Analysis

Upon excluding one participant with missing data on forgoing healthcare, we conducted a series of descriptive analyses. Chi-square tests were used to compare characteristics between those that did forgo healthcare, and those that did not. A multivariable logistic regression model separately assessed the above-mentioned risk factors for forgoing healthcare in general, adjusting for age and sex. This model was further stratified by the four most common reasons for forgoing healthcare. Analyses were done with R 4.0.3.

## Results

In total, we included 5,397 participants [55.4% women (0.4% intersex); mean age (SD): 51.7 (15.2) years)], of which 434 (8.0%) declared to have renounced healthcare since the start of the COVID-19 pandemic (Table 1). Among participants who reported forgoing healthcare, 63.6% were women, 25.8% were in a self-reported average-to-poor financial situation, and 22.4% reported an average or low health. The most common types of forgone healthcare were dental care (42.9%), appointments with a specialist (37.1%), general practitioner (21.7%), and surgical procedures (10.8%). Appointment cancellation was the most frequent reason for forgoing healthcare (53.9%), followed by fear of infection (35.3%), organizational issues (11.1%), and financial reasons (8.8%). 5.3% of participants who renounced healthcare reported doing so to avoid overloading the healthcare system (Table 1).

**Table 1.** Participants’ characteristics according to forgoing healthcare status, type of forgone healthcare and reason for forgoing healthcare - Specchio-COVID19 Study, Geneva, Switzerland.

|  | Forgone Healthcare |  |  | p-value |
| --- | --- | --- | --- | --- |
|  | N | No | Yes |  |
|  |  | N (%) | N (%) |  |
| **Total** | 5397 | 4963 (92.00) | 434 (8.00) |  |
| **Age (N=5397)** |  |  |  |  |
| 18-34 | 726 | 672 (92.56) | 54 (7.44) | 0.104 |
| 35-64 | 3408 | 3114 (91.37) | 294 (8.63) |  |
| 65+ | 1263 | 1177 (93.19) | 86 (6.81) |  |
| **Sex (N=5397)** |  |  |  |  |
| Men | 2386 | 2231 (93.50) | 155 (6.50) | 0.001 |
| Women | 2989 | 2713 (90.77) | 276 (9.23) |  |
| Intersex | 22 | 19 (86.36) | 3 (13.64) |  |
| **Country of birth (N=5397)** |  |  |  |  |
| Switzerland | 3217 | 2973 (92.42) | 244 (7.58) | 0.148 |
| Other | 2180 | 1990 (91.28) | 190 (8.72) |  |
| **Education (N=5394)** |  |  |  |  |
| Higher | 3425 | 3113 (90.89) | 312 (9.11) | 0.002 |
| Medium | 1676 | 1575 (93.97) | 101 (6.03) |  |
| Lower | 281 | 261 (92.88) | 20 (7.12) |  |
| Other | 12 | 11 (91.67) | 1 (8.33) |  |
| **Financial situation (N=5397)** |  |  |  |  |
| Very good | 2003 | 1866 (93.16) | 137 (6.84) | <0.001 |
| Good | 2122 | 1970 (92.84) | 152 (7.16) |  |
| Medium to poor | 845 | 733 (86.75) | 112 (13.25) |  |
| No Answer | 427 | 394 (92.27) | 33 (7.73) |  |
| **General health (N=5397)** |  |  |  |  |
| Good | 4801 | 4464 (92.98) | 337 (7.02) | <0.001 |
| Average | 530 | 449 (84.72) | 81 (15.28) |  |
| Poor | 66 | 50 (75.76) | 16 (24.24) |  |
| **Anti-SARS-CoV2-Serology (N=5396)** |  |  |  |  |
| Negative | 4563 | 4191 (91.85) | 372 (8.15) | 0.533 |
| Positive | 833 | 771 (92.56) | 62 (7.44) |  |
| **Type of Forgone Healthcare<sup>1</sup>** |  |  |  |  |
| Dental |  |  | 186 (42.9) |  |
| Specialist |  |  | 161 (37.1) |  |
| General |  |  | 94 (21.7) |  |
| Surgery |  |  | 47 (10.8) |  |
| Screening |  |  | 38 (8.8) |  |
| Physiotherapy |  |  | 26 (6.0) |  |
| Medical Device |  |  | 21 (4.8) |  |
| Drugs |  |  | 20 (4.6) |  |
| Alternative Medicine |  |  | 18 (4.1) |  |
| Outpatient Rehabilitation |  |  | 11 (2.5) |  |
| Health Center |  |  | 11 (2.5) |  |
| Other |  |  | 5 (1.2) |  |
| Psychologist |  |  | 4 (0.9) |  |
| Inpatient Rehabilitation |  |  | 3 (0.7) |  |
| Home Nursing |  |  | 3 (0.7) |  |
| **Reason for Forgoing Healthcare<sup>1</sup>** |  |  |  |  |
| Cancelled Appointment |  |  | 234 (53.9) |  |
| Fear of Infection |  |  | 153 (35.3) |  |
| Financial |  |  | 48 (11.1) |  |
| Organizational |  |  | 38 (8.8) |  |
| Conscious |  |  | 23 (5.3) |  |
| COVID Exposure |  |  | 21 (4.8) |  |
| Other |  |  | 15 (3.5) |  |
| Lethargy |  |  | 8 (1.8) |  |

Results are N (%). P-values are from Chi-Square Test. All variables are self-reported, except anti-SARS-CoV-2 serology (see Methods). <sup>1</sup> Participants could select multiple choices in the questionnaires.

The multivariable analysis showed that participants reporting an average (OR=2.55; 95% CI: 1.94-3.32) or poor health (OR=4.40; 95% CI: 2.40-7.68), or who were in a self-reported medium to poor financial situation (OR=2.04; 95% CI: 1.56-2.65) were more likely to forgo healthcare (Table 2). Participants with a medium education level were less likely than participants with a higher education level to forgo healthcare (OR=0.63; 95% CI: 0.50-0.79); the same pattern was observed for those with a low vs high education, though not significant. Country of birth and anti-SARS-CoV-2 serological status were not associated with forgoing healthcare in general.

**Table 2.** Association of sociodemographic risk factors, chronic disease and SARS-CoV-2 serology with forgoing healthcare, overall and for specific reasons - Specchio-COVID19 Study, Geneva, Switzerland.

|  | Any Reason | Fear of Infection | Organizational | Cancelled Appointment | Financial |
| --- | --- | --- | --- | --- | --- |
|  | OR (95% CI) | OR (95% CI) | OR (95% CI) | OR (95% CI) | OR (95% CI) |
| **Country of birth (N=5397)** |  |  |  |  |  |
| Switzerland | - | - | - | - | - |
| Other | 1.15 (0.94-1.40) | 1.53 (1.11-2.11)* | 2.04 (1.07-3.96)* | 1.09 (0.83-1.42) | 1.08 (0.60-1.92) |
| **Education (N=5395)** |  |  |  |  |  |
| Higher | - | - | - | - | - |
| Medium | 0.63 (0.50-0.79)** | 0.46 (0.29-0.69)** | 0.47 (0.19-1.01) | 0.60 (0.43-0.81)** | 0.79 (0.40-1.46) |
| Lower | 0.75 (0.45-1.17) | 1.04 (0.50-1.91) | 0.38 (0.02-1.80) | 0.76 (0.38-1.35) | 0.35 (0.02-1.64) |
| Other | 0.89 (0.05-4.61) | Undefined | Undefined | Undefined | Undefined |
| **Financial situation (N=5398)** |  |  |  |  |  |
| Very good | - | - | - | - | - |
| Good | 1.04 (0.82-1.32) | 0.88 (0.59-1.29) | 1.59 (0.68-3.98) | 0.98 (0.72-1.36) | 4.19 (1.08-27.52) |
| Medium to poor | 2.04 (1.56-2.65)** | 1.51 (0.97-2.33) | 2.72 (1.07-7.17)* | 1.85 (1.30-2.63)** | 42.20 (12.84-260.27)** |
| No Answer | 1.08 (0.72-1.59) | 1.05 (0.54-1.90) | 2.87 (0.92-8.42) | 1.00 (0.57-1.68) | 2.01 (0.09-21.15) |
| **General health (N=5398)** |  |  |  |  |  |
| Good | - | - | - | - | - |
| Average | 2.55 (1.94-3.32)** | 2.79 (1.82-4.17)** | 1.70 (0.57-4.09) | 2.96 (2.09-4.11)** | 1.02 (0.30-2.57) |
| Poor | 4.40 (2.40-7.68)** | 2.63 (0.79-6.57) | 2.54 (0.14-12.25) | 6.81 (3.49-12.41)** | Undefined |
| **Serology (N=5397)** |  |  |  |  |  |
| Negative | - | - | - | - | - |
| Positive | 0.89 (0.66-1.17) | 0.48 (0.26-0.82)* | 1.75 (0.80-3.55) | 0.93 (0.63-1.33) | 1.09 (0.49-2.18) |

Results are odds ratios (OR) and 95% confidence intervals (CI) from multivariable logistic regression, adjusted for age and sex.
\* indicates $p < 0.05$ ; \*\* indicates $p < 0.01$

When stratifying by reasons for forgoing healthcare, other patterns emerged (Table 2). Compared with participants born in Switzerland, those born elsewhere were more likely to forgo healthcare due to fear (OR=1.53; 95% CI: 1.11-2.11) and for organizational reasons (OR=2.04; 95% CI: 1.07-3.96). SARS-CoV-2 seropositive participants were less likely to forgo healthcare due to fear (OR=0.48; 95% CI: 0.26-0.82), whereas participants with an average reported health were more likely to forgo healthcare due to fear (OR=2.79; 95% CI: 1.82-4.17) and cancelled appointments (OR=2.96; 95% CI: 2.09-4.11) compared to those declaring a good health. On the opposite, participants with a medium education level were less likely to forgo healthcare due to cancelled appointments (OR=0.60; 95% CI: 0.43-0.81) or fear (OR=0.46; 95% CI: 0.29-0.69), compared to participants with a higher education level. When compared with the most financially privileged participants, individuals in a medium to poor financial situation were more likely to forgo healthcare for financial reasons (OR= 42.20; 95% CI: 12.84-260.27), organizational reasons (OR=2.72; 95% CI: 1.07-7.17) and cancelled appointment (OR=1.85; 95% CI: 1.30-2.63).

## Discussion

In a population-based sample of the adult population of Geneva, Switzerland, 8.0% of participants reported having forgone healthcare eight to ten months after the start of the COVID-19 pandemic, which contrasts with the much higher 41% reported by adults in the United States four to five months into the pandemic [6]. Since our study was conducted later into the pandemic, healthcare that was initially forgone may have been rescheduled, leading to fewer reports of forgone healthcare. This remains concerning as delayed healthcare can also lead to unfavorable health outcome [7,9]. Additionally, unlike the United States, healthcare coverage in Switzerland is universal [14], which may also explain part of this difference. As previously reported in Europe [1,4], we found that participants with an average or bad health, and those in a self-reported disadvantaged financial situation were more likely to forgo healthcare. Interestingly, around 5% of participants who renounced healthcare reported having renounced their healthcare appointments with the intention to prevent overburdening the system and to allow access for those with greater medical care needs. As this reason was not explicitly listed in the survey question but rather reported in the “other” option, the proportion of people choosing to forgo healthcare for altruistic reasons may be higher, especially concerning non-urgent healthcare. The most frequent type of forgone healthcare were dental, specialist, and general medicine, which reflects findings in the United States [3]. A study conducted before the pandemic among adults in Geneva found that 13.8% and 10.9% reported forgoing healthcare and dental care for economic reasons, respectively, which are a higher rate than in our sample [5,15]. The discrepancy could be attributed to how our survey question was interpreted; it defined forgoing healthcare since the beginning of the pandemic, but participants who would have renounced healthcare regardless may have not felt concerned.

One-third of participants who reported forgoing healthcare did so due to fear of SARS-CoV-2 infection. This reason was less common among SARS-CoV-2 seropositive participants; perhaps because they felt protected by the previous infection. Reverse causality is also possible, careless behavior increasing the risk of infection. Fear of being exposed to the virus in healthcare facilities is not exclusive to the COVID-19 pandemic; it has been well described in other infectious disease epidemics, including HIV and SARS [16]. Participants with a higher level of education were more likely to forgo healthcare due to fear; this was unexpected, given that this population also claimed to have fewer health concerns during the COVID-19 pandemic than those with a lower level of education [17,18]. However, avoiding healthcare centers could be considered as a COVID-19 preventive behavior, which seems to be more common among people with higher education levels [18]. Participants who reported average or poor health were more likely to renounce healthcare due to fear, probably because they felt more at-risk of severe COVID-19 outcomes [19]. A similar pattern was observed among participants born outside of Switzerland, which may reflect increased difficulties to navigate and access reliable information in a foreign healthcare system [20], especially during health crises. Clear and widespread messaging about the risks of forgoing needed healthcare are necessary to limit this phenomenon.

Over half of all forgone healthcare was a result of the healthcare facility cancelling appointments, which parallels findings in the United States [3]. More specifically, participants reporting an average or poor health were more likely to have a medical appointment cancelled. This could be attributed to those with health condition having more scheduled consultations that were cancelled when healthcare facilities reallocated resources for COVID-19 patients and may point to underlying gaps in both the capacity and accessibility of public health infrastructure in the wake of an epidemic. More research is needed to estimate how a decrease in the frequency of medical follow-up may affect people with existing health condition.

Many hospitals have reported between 23% to 49% decreased healthcare utilization during the COVID-19 pandemic [8]. Decreases in consultations with both general practitioners and physicians alike may have adverse health consequences, including complications and hospitalizations [21]. There is great concern that this decreased interaction with the healthcare system could also result in decreased access to routine childhood immunization services [22], and screening for comorbidities. The risk of forgoing healthcare can result in unfavorable health outcomes; a publication from the United States’ Urban Institute’s September 2020 Coronavirus Tracking Survey [3] illustrated that one in three adults who forwent or delayed healthcare during the COVID-19 pandemic had negative affects to their health, and ability to work or perform other daily activities. Another study found that 33% of the excess deaths observed between March and July 2020 in the United States was not attributed to COVID-19, whether due to undocumented SARS-CoV-2 infection or healthcare disruption [23]. In this context, hospitals and medical centers are increasingly using telemedicine-based services to improve patient care and provide consultations to patients who are unable to attend in-person appointments [24], though this aspect was not evaluated in the current study.

To our knowledge, this is among the few studies that present the type of renounced healthcare and reasons for forgoing it during the COVID-19 pandemic in a population-based sample [6]. The cross-sectional design is a limitation to our findings, as it prevented us from examining changes in forgoing healthcare patterns since the start of the COVID-19 pandemic. The baseline questionnaire also limited the scope of our analysis; we were unable to assess if forgone healthcare had any negative health impacts, nor make a distinction between renounced or delayed healthcare. Forgoing healthcare may have been underestimated in our study due to social desirability recall bias and because of the potential underrepresentation of underprivileged populations.

## Conclusion

In conclusion, in the Geneva adult population, only 8.0% of people reported forgoing healthcare since the beginning of the COVID-19 pandemic, 8 to 10 months after the first lockdown. People with a disadvantaged financial situation, and those reporting an average or poor health were more likely to forgo healthcare. Forgoing healthcare has long-standing repercussions for morbidity and mortality, and these results highlight the risk of widening gaps between privileged and vulnerable populations. Further exploration of underlying reasons for delaying or avoiding healthcare altogether are needed for epidemic-preparedness planning. More research also is needed to better understand the implications of forgoing healthcare for individuals with health condition. In the interim, increasing accessibility of medical and telehealth services, especially for vulnerable groups, might help prevent delay of needed care.

## Data Availability

Our data are accessible to researchers upon reasonable request for data sharing to the corresponding author.

## Acknowledgment

We are grateful to the staff of the Unit of Population Epidemiology of the University hospital of Geneva, Division of Primary Care Medicine, as well as to all the participants whose contributions were invaluable to the study.

